# Patterns of sacral dysmorphism in pelvic CT scans at a national referral hospital in Kenya

**DOI:** 10.1101/2023.12.20.23300320

**Authors:** Valentine B Nyang’au, Fred Sitati, Edward Gakuya, Oline Amunga

**Affiliations:** Department of Orthopedics, University Health Services, University of Nairobi, Kenya; Department of Surgery, Orthopaedic Unit, University of Nairobi, Kenya

**Keywords:** sacral dysmorphism, sacral dysmorphism score, pelvic CT (computed tomography), sacroiliac screws, S1, S2

## Abstract

**Background:** Sacral dysmorphism refers to morphological variations found in the first two sacral segments that limit the safe placement of percutaneous sacral iliac screws. The prevalence is documented in European, North American and some Asian populations. However, studies within the African population including Kenya are lacking. The aim of the study was to describe the patterns of sacral dysmorphism in pelvic computerized tomography (CT) scans at a national referral hospital in Kenya.

**Methods:** A cross-sectional study carried out at the Radiology Department, Kenyatta National Hospital from March 2020 to March 2021 involving the radiographic evaluation of 293 stored abdominal pelvic CT scans of patients. Sacral dysmorphism was identified based on the sacral dysmorphism score >70 and the presence of any of the six morphological features of sacral dysmorphism.

**Results:** A sacral dysmorphism score of more than 70 was found to in 64% of the population. The prevalence of dysmorphic sacra (based on the presence of at least one qualitative feature) was 100%. The most prevalent feature of sacral dysmorphism was the lack of recession of the S1 segment (82%) followed by an unfused sacral segment (76%). Of note is that some qualitative features of sacral dysmorphism were protective against a high sacral dysmorphism score. There was no statistical significance of gender in sacral dysmorphism.

**Conclusion:** There is a high prevalence of sacral dysmorphism score in this population. There is a need for further studies to revisit the concept of dysmorphic sacra based on the presence of at least one qualitative feature as all our participants had at least one feature.

## INTRODUCTION

The term *“dysmorphic or dysplastic”* sacra refers to abnormal variations in the upper sacral segments (S1 and S2) that may create a narrow or more angulated intraosseous corridor that would limit the safe passage of percutaneous sacral iliac screws. These set of features were first described on review of plain pelvic radiographs(1). These features include:-lack of sacral fusion, irregular sacral foramina, lack of collinearity of the iliac crest, presence of mammillary bodies, acute alar slopes and presence of a tongue in groove phenomenon. The determination of sacral dysmorphism is done based on the presence of these descriptive features with some studies defining sacral dysmorphism as the presence of all of the descriptive features while others base it on the presence of any of the above descriptive features. This has led to increased research on quantitative methods to determine sacral dysmorphism.

Percutaneous sacral iliac screws is the current gold standard in fixing posterior pelvic disruption(2). These screws provide effective stability and haemorrhage control and are superior to traditional open approaches which are associated with significant morbidity and mortality. It is important for the surgeon to be conversant with the identification of sacral dysmorphism to make relevant pre-operative plans to ensure safe placement of these screws.

There is a significant knowledge gap globally on sacral dysmorphism. Qualitative methods of identifying sacral dysmorphism are subjective; newer quantitative techniques such as the sacral dysmorphism score are being described(3). The prevalence of sacral dysmorphism has been found to be high ranging from 40-70 percent; these studies have mostly been done in European(4), North American(3) and Asian populations(5). There was no data in the African set up.

The aim of the study was to describe the patterns of sacral dysmorphism in pelvic computerized tomography (CT) scans at a national referral hospital in Kenya and this goal was achieved.

## MATERIALS AND METHODS

This was a cross-sectional study. It involved radiographic evaluation of 293 lumbosacral and pelvic helical CT scans (T12-L5). We included scans of patients aged between 18 and 70 years and had visualization of the lowest rib-bearing vertebra up to the lesser trochanter. Scans with unstable pelvic ring injury, implants obscuring the lumbosacral junction, sacral tumors or infection and lumbar scoliosis of more than 20 degrees, or spina bifida were excluded. Any scan that did not meet the above criteria was replaced by selecting the next randomized participant. This study was conducted at Kenyatta National Hospital (KNH), a national referral hospital, Kenya Essential Package for Health (KEPH) level six, in Nairobi County. The institution is a tertiary referral facility with specialized radiologic and orthopaedic services and staff. The study used digital repositories of stored pelvic CT scan images in the Radiology Department at KNH for a period of 13 months from March 2020 to March 2021. A simple random sampling technique was used in this study because it offered an equal chance of selection in the study population. The study population was tabulated on Microsoft Excel and randomized using the random function.

The direction of the scanning was cranio caudal; no contrast was given, and the respiratory phase was suspended. Patients lay supine with hips and knee joints extended. Original axial images were presented in different presets including bone window and converted to multiplanar reconstruction (MPR) and three-dimensional rendering. The CT was conducted with a NEOSOFT–64 slice scanner. Images taken were reformatted to 2mm thickness to better define features.

The population size, *N*, was 1200 pelvic CT scans, these are from the CT scan repository in KNH for the duration of the study period. The anticipated frequency of sacral dysmorphia*, p* was 40%(3). The confidence interval was 95% and the design effect (DEFF) was one. The value of standard normal distribution corresponding to a 95 percent significant level, Z^2^_1-α_. Finally, the study had a powered^2^, of 80 percent. A sample size of 283 was calculated using the OpenEpi online sample size calculator (*Epi Calc*, 2021.)Using the sampling equation for a finite population: *sample size n = [DEFF*Np (1-p)]/ [(d^2^/Z^2^ *(N-1) +p*(1-p)]*.

In our final data collection, 303 CT scans were sampled, 10 records were omitted during data cleaning; as they has spurious data and finally 293 records were analysed; this figure ensured our study was adequately powered. Outliers, especially for sagittal angle of more than70 and below 10 degrees were dropped since they are not physiologically plausible.

Excluded data was not analysed. There was no missing data since the electronic data extraction tool had mandatory questions and skip patterns inbuilt in it based on the inclusion and exclusion criteria.

The study population was tabulated on Microsoft Excel and randomized using the random function.

### Sacral Dysmorphism Score

To calculate the sacral Dysmorphism Score, each CT scan was reformatted along the axis of the sacrum to obtain cross-sectional imaging of the S1 and S2 sacral osseous corridor, or *‘‘safe zone’’* for iliosacral screw placement(7). For standardization and reproducibility; the scans were reformatted in true coronal and axial planes by defining the sagittal axis as the posterior border of the sacrum and the axial reformats along the upper-end plate of the S2 vertebrae(8).Coronal angulation was measured by an angle formed by a line drawn along the axis of the osseous corridor and a line connecting the top of the iliac crests(7). The top of the iliac crest was found by correlating the axial and sagittal views on multi-planar reconstruction. Axial angulation was measured as the angle formed by a line drawn on the axis of the osseous corridor and a line connecting the posterior iliac spines. The tip of the posterior superior iliac spine was found by correlating the axial and sagittal views on MPR reconstruction. The sacral dysmorphism score = (first sacral coronal angle) + 2 (first sacral axial angle)(7).The outcome variable, sacral dysmorphism, was defined as a dysmorphic score of more than 70 on the quantitative analysis(3).

### Sacral dysmorphism based on qualitative characteristics

To evaluate the qualitative characteristics of sacral dysmorphism, we created pelvic outlet reconstruction in which neutral horizontal rotation was corrected by aligning lumbar spinous processes with the symphysis pubis. The vertical rotation was adjusted to align the superior cortex of the pubis with the second sacral segment body(7). Each outlet reconstruction was reviewed and the presence or absence of each of the radiographic qualitative characteristics of sacral dysmorphism that included: an upper sacral segment not recessed in the pelvis; the presence of mammillary processes; an acute alar slope; a residual disc between the first and second sacral segments; and noncircular upper sacral neural foramina determined. In addition, axial CT scans were reviewed for the presence or absence of a *‘‘tongue-in-groove’’* sacroiliac morphology(1). The definition of sacral dysmorphism based on the number of dysmorphic features has been controversial in our case we assessed dysmorphism as the presence of any of the above features. This is because each of these features has been associated with narrowing the safe corridor for passage of sacral iliac screws(1).

### Ethical approval

The study was approved by Kenyatta National Hospital-University of Nairobi Ethics and Research Committee (KNH-UON ERC) approval number KNH-ERC/A/9 as required by the Department of Surgery, Orthopaedic Unit Faculty of Health Sciences, University of Nairobi. The study was also approved by the National Commission for Science Technology and Innovation NACOSTI reference NACOSTI/P/22/15319.

### Data analysis

We performed univariate analysis using Stata 14 ® to produce proportions, tables, charts, and graphs. In addition, bivariate analysis was done as a measure of association with sacral dysmorphism score as the outcome variable and other exposure variable such as sex, age and each of the six qualitative features.

## RESULTS

Out of the 293 randomly selected CT scans, the majority were male (53%).The median age was 44years with an inter quartile range of 34-55years. Sacral dysmorphism score of more than 70 was found in 64% of the study participants, with it being more prevalent among males at 69% compared to females respectively. The prevalence of sacral dysmorphism based on presence of at least one qualitative feature was 100%.

The most prevalent dysmorphic feature in the normal population was tongue in groove (73%) and acute alar slope (59%) respectively, whereas the most prevalent features of sacral dysmorphism in the dysmorphic population was the unfused sacral segment (67%) and lack of recession of the s1segment(66%). See Figure 3.

**Figure 1:**
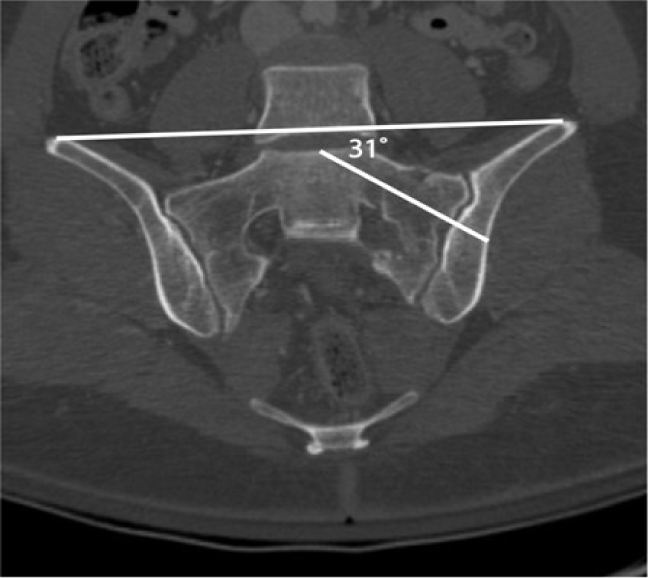
Coronal angulation was measured as the angle subtended by a line drawn along the axis of the osseous corridor and a line connecting the top of the iliac crests(7)

**Figure 2:**
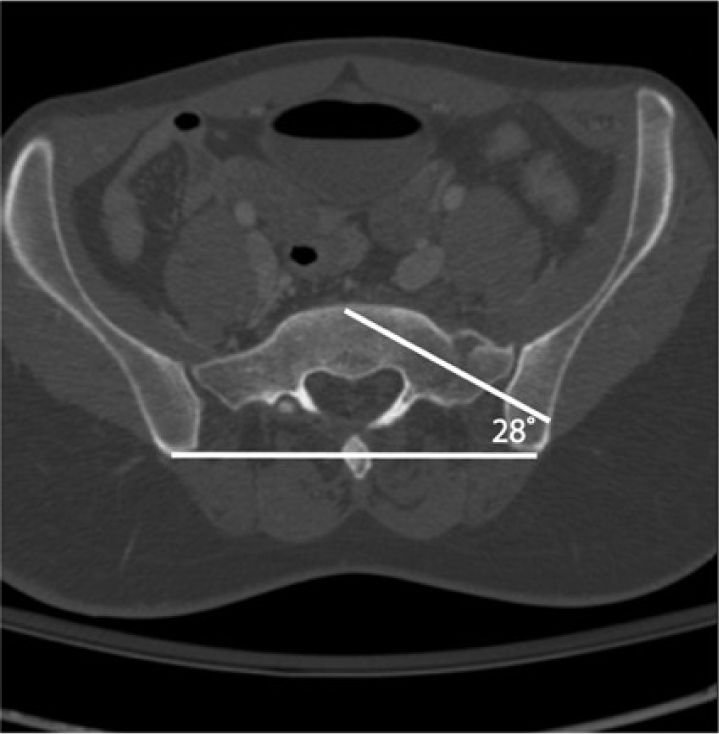
Axial angulation was measured as the angle formed by a line drawn on the axis of the osseous corridor and a line connecting the posterior iliac spines(7)

**Figure 3:**
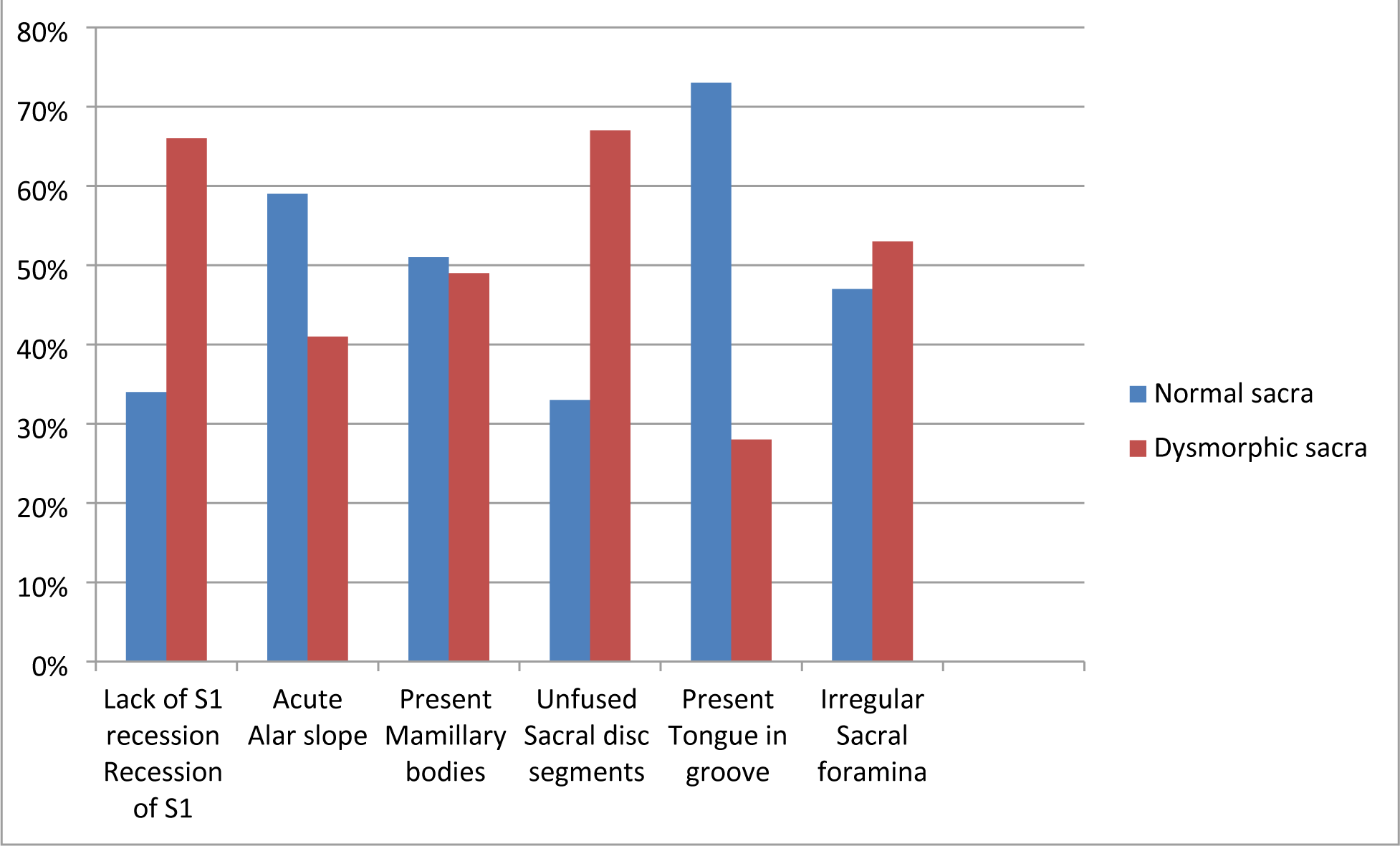
Proportion of radiological features of sacral dysmorphism in the normal sacra and dysmorphic sacra as categorized by the sacral dysmorphism score.

Sacral dysmorphism score was significantly higher in males compared to females p=0.0459. We found that the prevalence of four radiological features of sacral dysmorphism were statistically significant p =<0.01 compared to the normal sacral features these are non acute slope, absent mammillary bodies, absent tongue in groove and regular sacral foramen. These four features were more prevalent in the dysmorphic sacra as compared to the normal sacra. See Table 1.

**Table 1:** Comparative analysis of sacral dysmorphism score by sex and radiological features.

| Variable |  | Sacral dysmorphism score |  | Total | p-value |
| --- | --- | --- | --- | --- | --- |
|  |  | Normal sacra | Dysmorphic sacra |  |  |
| Sex | Female | 56 (41%) | 81 (59%) | 137 | **0.0459** |
|  | Male | 49 (31%) | 107 (69%) | 156 |  |
| Recession of S1 | Lack of S1 recession | 81 (34%) | 159 (66%) | 240 | 0.0565 |
|  | Recessed S1 | 24 (45%) | 29 (55%) | 53 |  |
| Alar slope | Not acute | 58 (27%) | 155 (73%) | 213 | **<0.0001** |
|  | Acute | 47 (59%) | 33 (41%) | 80 |  |
| Mamillary bodies | Absent | 67 (31%) | 151 (69%) | 218 | **0.0010** |
|  | Present | 38 (51%) | 37 (49%) | 75 |  |
| Sacral disc segments | Unfused | 75 (33%) | 149 (67%) | 224 | 0.0650 |
|  | Fused | 30 (43%) | 39 (57%) | 69 |  |
| Tongue in groove | Absent | 47 (22%) | 166 (78%) | 213 | **<0.0001** |
|  | Present | 58 (73%) | 22 (28%) | 80 |  |
| Sacral foramina | Regular | 54 (29%) | 130 (71%) | 184 | **0.0013** |
|  | Irregular | 51 (47%) | 58 (53%) | 109 |  |

**\*Two-sample test of proportions- z test was used to test if the difference between the proportions was** **significant**

Crudes odds ratio was carried out as a measure of association between the various radiological features of sacral dysmorphism and the sacral dysmorphism score. We found significant results P<0.01 in four radiological features; acute alar slope, mammillary bodies, tongue in groove formation and sacral foramina. Scans that had a non acute alar slope, absent mammillary bodies, absent tongue in groove malformation and regular sacral foramina were more likely to have a sacral dysmorphism score of >70. See Table 2.

**Table 2:** Bivariable analysis of factors associated with sacral dysmorphism.

| Variable |  |  | Sacral dysmorphism score |  | Total | Odds ratio | 95% CI | p-value |
| --- | --- | --- | --- | --- | --- | --- | --- | --- |
|  |  |  | Normal sacra | Dysmorphic sacra |  |  |  |  |
| Sex | Female |  | 56 (41%) | 81 (59%) | 137 | 1.5 | 0.9-2.5 | 0.0918 |
|  | Male |  | 49 (31%) | 107 (69%) | 156 |  |  |  |
| Recession of S1 | Lack of S1 recession |  | 81 (34%) | 159 (66%) | 240 | 0.6 | 0.3-1.2 | 0.1130 |
|  | Recessed S1 |  | 24 (45%) | 29 (55%) | 53 |  |  |  |
| Alar slope | Not acute |  | 58 (27%) | 155 (73%) | 213 | 0.3 | 0.1-0.5 | <0.0001 |
|  | Acute |  | 47 (59%) | 33 (41%) | 80 |  |  |  |
| Mamillary bodies | Absent |  | 67 (31%) | 151 (69%) | 218 | 0.4 | 0.2-0.8 | 0.0019 |
|  | Present |  | 38 (51%) | 37 (49%) | 75 |  |  |  |
| Sacral disc segments | Unfused |  | 75 (33%) | 149 (67%) | 224 | 0.7 | 0.4-1.2 | 0.1300 |
|  | Fused |  | 30 (43%) | 39 (57%) | 69 |  |  |  |
| Tongue in groove | Absent |  | 47 (22%) | 166 (78%) | 213 | 0.1 | 0.1-0.2 | <0.0001 |
|  | Present |  | 58 (73%) | 22 (28%) | 80 |  |  |  |
| Sacral foramina | Regular |  | 54 (29%) | 130 (71%) | 184 | 0.5 | 0.3-0.8 | 0.0026 |
|  | Irregular |  | 51 (47%) | 58 (53%) | 109 |  |  |  |

## DISCUSSION

Identification of sacral dysmorphism is important for pre-operative planning when placing percutaneous sacral iliac screws(4).Percutaneous sacral iliac screws are likely to be misplaced in cases of dysmorphism(9). Misplaced screws may cause long term morbidity due to injury of close neurovascular structures. However, the definition of sacral dysmorphism is not standardized; with the question of how many radiological features does it take to label a sacrum as dysmorphic not answered(4). Studies carried out in the African population are to the best of our knowledge absent with most done in European(10), North American(9) and Asian populations(11).

In our study, the prevalence of sacral dysmorphism based on the sacral dysmorphism score was 64 % whereas the prevalence of sacral dysmorphism based on the presence of at least one feature was 100%. Sacral dysmorphism score of more than 70 in the original study by Kaiser et al was found to be 41% in the American population(3), 84.7% in the Iranian population(12) and a mean of 47.4± 15.8 in the German population(10).

The most prevalent dysmorphic features were the presence of unfused sacral segments 67% and lack of recession of the s1 segments at 66%. This was similar to other studies where presence of unfused sacral segments was the most prevalent dysmorphic feature 70% in a German based study (4); 71% in the Iranian study(12) and 50% in the Mugla region in Turkey(13). As evidenced by these studies, un fused sacral segments may be used as a screening tool for sacral dysmorphism as it has been largely associated with the dysmorphic sacrum (sacral dysmorphism score >70) unlike other features which have been highly variable. The sacral discs are separated in childhood with the lower segments S3-S4and S4-S5fusing in adolescence and the upper segments by the 3^rd^ decade of life(14). This may suggest a relationship with age, however in our study no age association was found similar findings to other studies(4).

It is important to note that the dysmorphic features were also present in the normal sacrum(sacral dysmorphism score< 70).These findings are similar to other studies, where dysmorphic features are still found within the normal population(12).Some of these features such as mammillary bodies, an acute alar slope, the presence of a tongue in groove formation and irregular sacral foramina were found to be more prevalent in the normal population. This suggests that these features may form a normal anatomical variant in our population in our population. The finding on the tongue in groove feature as seen on axial views of the CT scan has been found to negatively correlate with sacral dysmorphism, similar to our study findings(3).We found no evidence in gender differences in sacral dysmorphism based on the sacral dysmorphism score. This is similar to other recent studies which noted no statistical gender differences in sacral dysmorphism occurrence(3).

The high prevalence of sacral dysmorphism based on qualitative features within our population may suggest that these features could be normal anatomical variants. These features alone may not be objective in determining the presence of dysmorphism. We found that all the pelvises had at least one dysmorphic characteristic based on the initial descriptions of sacral dysmorphism (15). We found studies that showed high rates of dysmorphism, such us up to 85%(4); however none showed a 100% prevalence where each scan had at least one dysmorphic feature. These are suggestive of a normal anatomical variant in our population.

Our study is to the best of our knowledge, the first in an African population. This was limiting to our findings as we could not compare local results to other studies carried out within the African continent. The role of ethnicity in the prevalence and patterns of sacral dysmorphism cannot be ignored and presents an exciting front for future research.

## CONCLUSION

Sacral dysmorphism is highly prevalent within our study population. Care should be taken in screening patients for sacral dysmorphism as part of the pre-operative planning for sacral iliac screw fixation. Descriptive or qualitative features of sacral dysmorphism do not allow for a reliable way of predetermining sacral dysmorphism. Quantitative measures such as the sacral dysmorphism score offer a more objective way to evaluate sacral dysmorphism. This is relevant for both orthopaedic surgeons and radiologists.

## Data Availability

All relevant data are within the manuscript and its Supporting Information files.

## ACKNOWLEDGMENTS

We wish to acknowledge the training and support from the University of Nairobi’s Building Capacity for Writing Scientific Manuscripts (UANDISHI) Program at the Faculty of Health Sciences. This work was funded in part through the ADVANCE program at IAVI. This work is made possible by the support of the American People through the U.S. President’s Emergency Plan for AIDS Relief (PEPFAR) through United States Agency for International Development (USAID). *The contents of this studyare the sole responsibility of the authorsand do not necessarily reflect the views of PEPFAR, USAID, or the United States Government*.

